# Instrumental variable analysis: choice of control variables is critical and can lead to biased results

**DOI:** 10.1101/2024.07.11.24310262

**Authors:** Fergus Hamilton, Todd C Lee, Guillaume Butler-Laporte

## Abstract

Instrumental variable (IV) analysis is a widely used technique in econometrics to estimate causal effects in the presence of confounding. A recent application of this technique was used in a high-profile analysis in *JAMA Internal Medicine* to estimate the effect of cefepime, a broad-spectrum antibiotic, on mortality in severe infection. There has been ongoing concern that piperacillin-tazobactam, another broad-spectrum antibiotic with greater anaerobic activity might be inferior to cefepime, however this has not been shown in randomized controlled trials. The authors used an international shortage of piperacillin-tazobactam as an instrument, as during this shortage period, cefepime was used as an alternative. The authors report a strong mortality effect (5% absolute increase) with piperacillin-tazobactam. In this paper, we closely examine this estimate and find it is likely conditional on inclusion of a control variable (metronidazole usage). Inclusion of this variable is highly likely to lead to collider bias, which we show via simulation. We then generate estimates unadjusted for metronidazole which are much closer to the null and may represent residual confounding or confounding by indication. We highlight the ongoing challenge of collider bias in empirical IV analyses and the potential for large biases to occur. We finally suggest the authors consider including these unadjusted estimates in their manuscript, as the large increase in mortality reported with piperacillin-tazobactam is unlikely to be true.

## Introduction

In a recent issue of *JAMA Internal Medicine*, Chanderraj et al^1^ provide an innovative analysis attempting to use a shortage of a widely used broad-spectrum antimicrobial, piperacillin-tazobactam (TZP), to conduct a comparative effectiveness study versus cefepime (CPM), an agent used as a replacement during this period which has a similar spectrum of activity except less anaerobic activity. Their instrumental variable (IV) analysis^1^ utilises the ‘natural experiment’ of substitution of TZP with CPM over this period in a manner which is, in principle, unrelated to other confounders. They hypothesize that the additional anti-anaerobic activity of TZP might lead to increased mortality via effects on the microbiome or resistome. Their primary analysis using a two-stage-least-squares (TSLS) IV analysis found that TZP use was associated with a 5% absolute increase in mortality (95% CI 1.8-8.1%, p = 0.002). This was reported as:

> *“administration of piperacillin-tazobactam was associated with higher mortality and increased duration of organ dysfunction compared with cefepime”*

This is in stark contrast to recent randomized controlled trial data directly comparing the two drugs which do not suggest such a benefit.^2^ Chanderraj et al’s result was widely discussed (Altmetric score 457 on 2^nd^ July) and highlighted in *NEJM Journal Watch*^3^, which concluded:

> *“Therefore, we can infer that among severely ill patients with undifferentiated sepsis, treatment with cefepime rather than PTZ [piperacillin-tazobactam] is associated with an absolute mortality reduction at 90 days*.*”*

Interestingly, despite a marked reduction in piperacillin-tazobactam and increase in cefepime usage during the shortage period, there was no independent effect of the shortage period on mortality (20.8% outside shortage, 19.8% within shortage, p = 0.31). This is unusual and raises a concern. In general, TSLS estimates are more imprecise and have larger standard errors than the independent IV-outcome analysis reflecting the uncertainty involved in the twostep process.^4^ Less formally, there are few plausible explanations whereby all three of the below hold:

1. The instrument (shortage period) strongly influences the exposure (cefepime usage)
2. The instrument (shortage period) has no influence on the outcome (mortality)
3. The exposure (cefepime usage) has a very strong effect on the outcome (mortality)

In this short analysis, we explored potential reasons for the difference between the TSLS estimate (suggesting strong evidence of benefit) and the raw analysis (suggesting no effect) and postulate that this has occurred due to erroneous inclusion of control variables in the analysis. Control variables are often included to increase precision of TSLS estimation but should only be included under certain conditions and when they are upstream of both the instrument and the outcome.^5^ For example, an analysis using state or country level cigarette taxation as an instrument for the outcome of lung cancer might include socio-economic status of the participant as a control variable to increase precision. Or similarly, analyses using genetic variants as instruments might include age of participants as control variables, as this cannot be plausible caused by genetics, but may influence the outcome.

However, the inclusion of control variables that are downstream of the IV should generally be avoided^5^, and can be considered as being similar to inappropriately adjusting for post-randomisation variables in a randomised trial.^6^ In the presented analyses, the authors adjust for a number of demographic variables but also control for metronidazole usage in both stages of the regression. This is despite the authors reporting that the shortage period strongly influences metronidazole prescription (eFigure 2 in the original publication).

This is inappropriate and has the potential to lead to collider bias^7^ if unmeasured confounding affects both MTZ usage and mortality which is probable, as metronidazole usage is a marker of sickness (particularly when cefepime is used, as metronidazole is often co-prescribed for intra-abdominal infection where mortality is high). This collider bias induces an association between the IV and confounders (e.g. disease severity) of the association between the exposure (cefepime usage) and outcome (mortality). This directly contravenes a core assumption of IV analyses.

We first show via simulation why the addition of metronidazole into the IV analysis could lead to bias. We then present our own analysis using summary data from the trial to estimate the causal effect unadjusted for control variables.

## Methods

### Simulation study

For our simulation study, we aimed to match the potential real-life causal data-generating process, although we recognise such a simulation is an oversimplification, and the major value is in demonstrating the potential collider bias. We generated 5 simulated variables, reflecting the key components: Z (the shortage period, representing the instrument), X (the exposure, use of cefepime), M (metronidazole usage), Y (the outcome, mortality), U (unmeasured confounding). We set up the relationship between these variables as per **Figure 1**.

**Figure 1:**
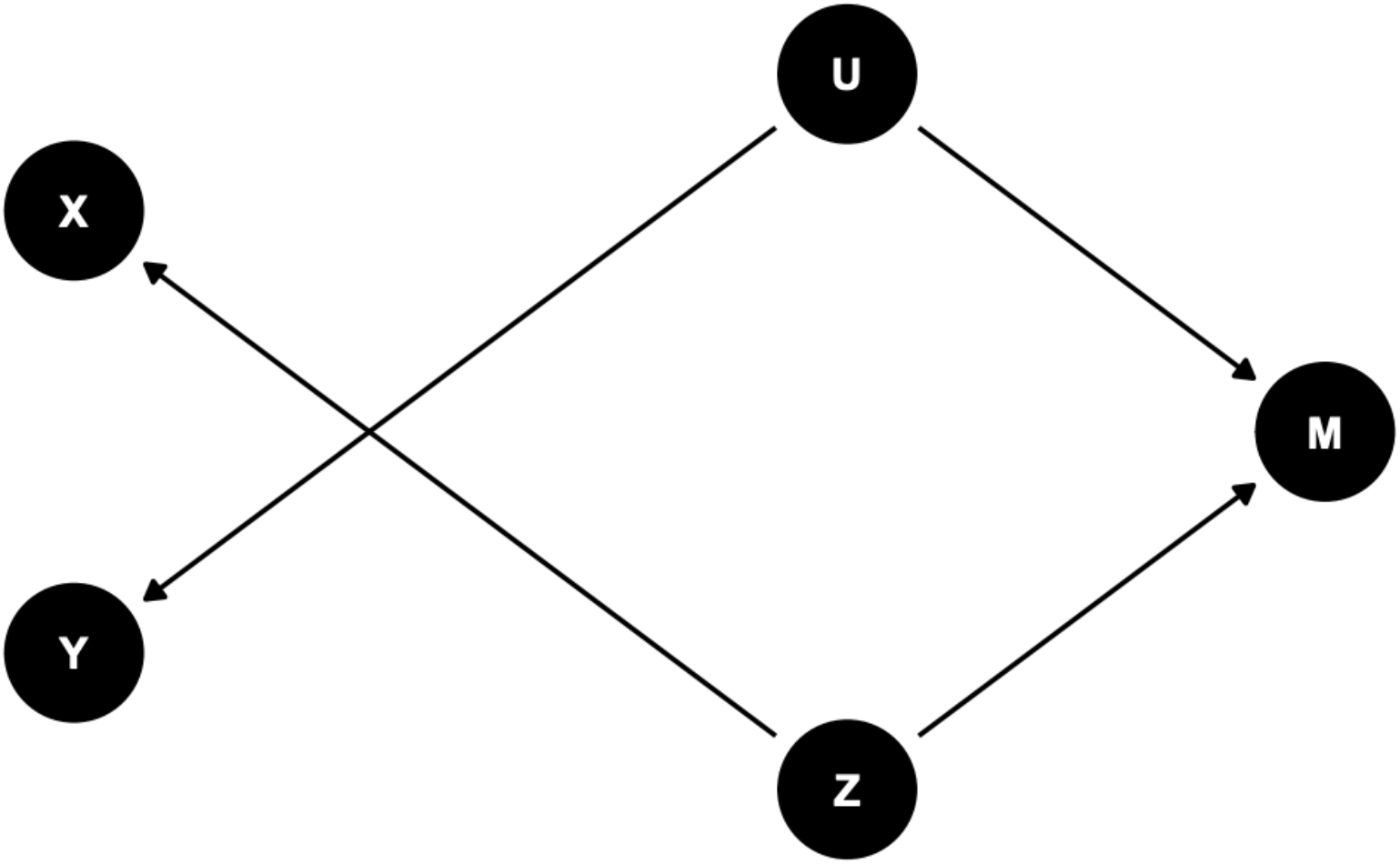
A directed acyclic graph (DAG) describing our simulation. Each point represents a variable, while each arrow represents the simulated effect direction.

In this situation, we have a strong relationship between Z and X (as expected, the shortage period led to a large increase in cefepime prescribing), but we also have a strong relationship between Z and M (as was reported in their eFigure2). We also simulated no causal effect of the exposure on the outcome (i.e. the prescription of cefepime has no effect on mortality). This can be seen by the lack of an arrow between X and Y. Finally, we simulated unmeasured confounding (U) which influences both mortality and metronidazole usage. This variable represents the fact that metronidazole usage is influenced by other factors that themselves associate with mortality (e.g. UTI (no metronidazole, lower mortality) vs. intrabdominal or undifferentiated sepsis (metronidazole, higher mortality)). We can strongly suspect this is the case, as metronidazole users had a strong marginal association with mortality in the reported logistic regression analysis (OR 1.54, 95% CI 1.32, 1.8, data from eTable 6 in the original publication^1^).

Simulated variables were generated in R for 5,000 participants, using the *tidyverse* package, and IV analysis was performed using the AER package to generate estimates. Further technical details and statistical code are provided in **Supplementary Note 1**. Of note, we can also simulate the equally plausible situation where M is entirely downstream of X, rather than Z (as the decision to give metronidazole is made after the decision to give piperacillin-tazobactam, and there is no independent effect of Z onto M. This does not fundamentally alter results but is reported in Supplementary Note 1 for completeness.

The important detail in this simulation is that we should not have any evidence of an effect in IV analyses; there is no relationship between the exposure and the outcome, and evidence of a relationship is a sign of bias.

#### Generation of summary level estimates from the original manuscript

Although estimates in IV analyses are usually made using individual level data, it is possible to generate estimates using summary level data. This is commonly performed in Two-Sample Mendelian randomisation, where only the summary effects of a genetic variant on an exposure and outcome are required.^8^

We can therefore estimate, accepting a degree of error, a TSLS estimate that is unadjusted for the control variables in Chanderraj et al by simply calculating the IV-outcome estimate, and dividing this by the IV-exposure estimate (the Wald ratio). The standard error of this estimate can be approximated by dividing the standard error of the IV-outcome estimate by the IV-exposure estimate. We calculate this (full details in the **Supplementary Note 2**) and compare this with the presented analyses.

#### Ethics

This analysis is a re-analysis of presented work and requires no ethical approval

#### Funding

FH’s time was funded by the NIHR Clinical Lectureship scheme.

## Results

### Simulation study

In our simulation study, we aimed to closely replicate the Chanderraj et al’s analysis.^1^ In the presented paper, they report that the IV strongly influences metronidazole (M) usage, alongside having a large effect on the exposure (X). In our simulation, we have no causal effect of the exposure on the outcome, so when analyses are run unadjusted for metronidazole, IV estimates are null (**Figure 2**).

**Figure 2:**
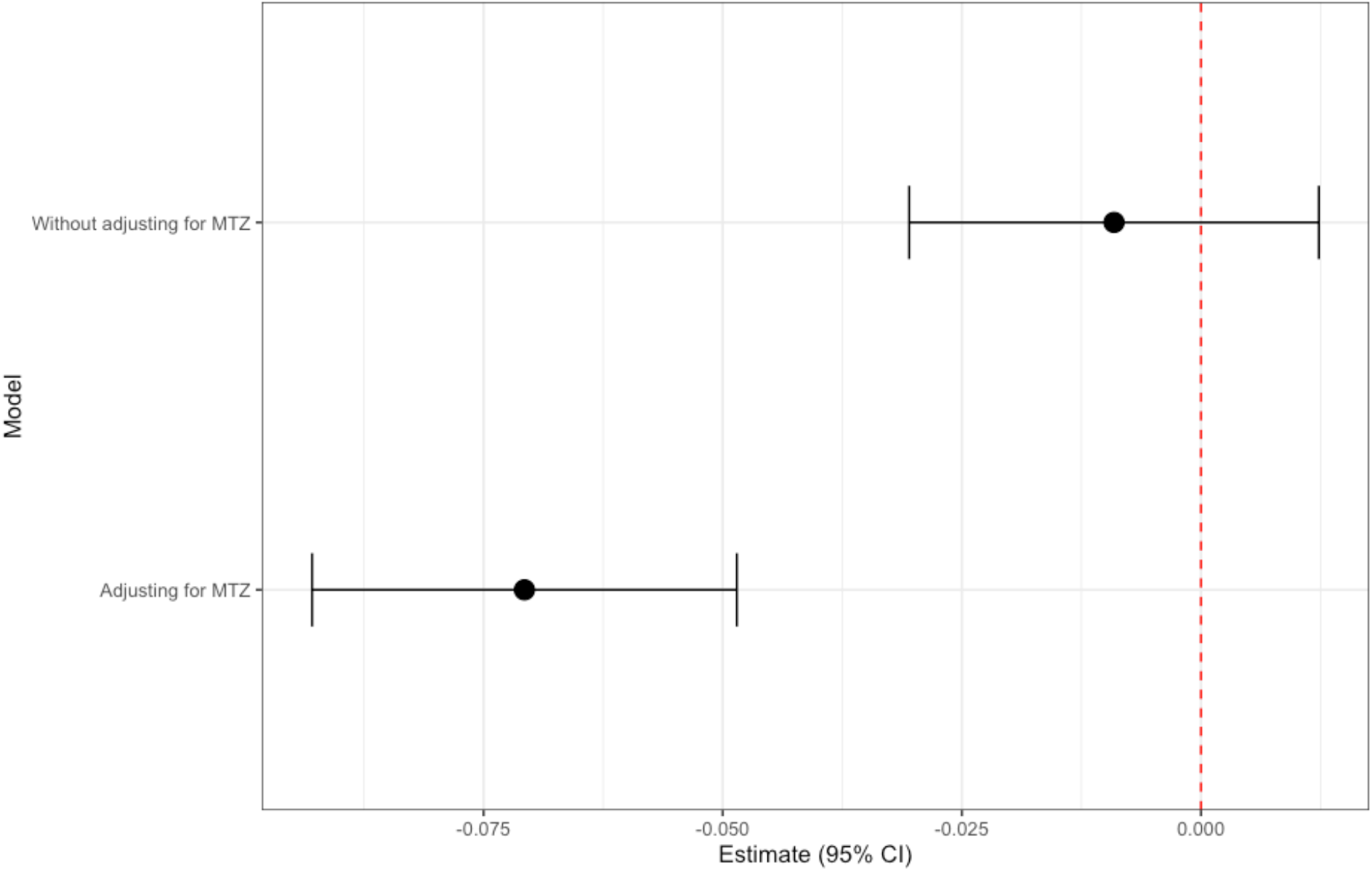
Simulated estimates of the effect of adjusting for metronidazole on an outcome using the causal model above. In the unadjusted model, causal estimates are close to the null, and are unbiased. In the adjusted model, estimates are biased and show a causal effect of X (“cefepime”) on the outcome.

However, when we include metronidazole in the model we induce a bias, and we get strong evidence of a protective effect of cefepime. This occurs despite there being no causal effect of cefepime on the outcome. The reasons for this counterintuitive result are worth discussing: the bias occurs because we have opened a path between the unmeasured confounding (U) and the outcome. Essentially, we bring back most of the bias that IV analyses are meant to protect against (unmeasured confounding). Consider that the patient only gets metronidazole during the shortage if the clinician thinks the patient is sick enough to require coverage for anaerobes which cefepime itself does not provide. By adjusting for metronidazole, we induce a negative relationship between patient sickness and the instrument that then biases the estimate on mortality.

This bias occurs in the same way when performing analyses stratified by metronidazole usage. Because metronidazole usage associates with mortality, in both metronidazole users and non-users, there is a negative relationship induced between the instrument and mortality which leads to biased estimates. Other examples of similar collider biases have been widely described in COVID-19 and are likely to explain (some) of the ‘obesity paradox’, the paradoxical relationship between increased BMI and better critical disease outcomes in severe infection.^9^ It is important to note that this bias also occurs if the causal path we present is incorrect, and metronidazole usage is entirely downstream of piperacillin-tazobactam usage, and the IV has no direct effect on this (**Supplementary Note 1**).

In summary, our simulation study shows that in the presence of unmeasured confounding of metronidazole usage and mortality, adjusting for metronidazole will induce bias in estimates of the effect of cefepime on mortality.

#### Generation of new estimates

Although our simulation study provided support for potential bias, to test this we needed to generate estimates unadjusted for metronidazole. We attempted to use the reported summary level data and estimated the causal effect of cefepime on mortality.

Details of this calculation are provided in **Supplementary Note 2** but can be summarised as calculating the effect of the shortage on piperacillin-tazobactam usage (IV-exposure effect, -0.659) and the effect of the shortage on mortality (IV-outcome effect, -0.0650) using logistic regression, and using the Wald ratio to generate the IV estimate of -0.065/-0.659, which corresponds to a log odds ratio of 0.098, and an odds ratio of 1.10, 95% CI 0.91-1.33. Formulas for standard error are in the S**upplementary Note 2**. The absolute change in mortality is then calculated using the odds ratio and baseline mortality.

Our final estimate is an absolute risk increase of 2.2% with piperacillin-tazobactam usage (95% CI -1.8% to 7.0%), weaker and more imprecisely estimated than their primary estimate (5%; 95% CI 1.8-8.1%, **Figure 3**). This does not provide compelling evidence of a strong effect of superiority of cefepime. We recognise the limitations of this analysis, but are unable to do more detailed analysis without access to individual level data.

**Figure 3:**
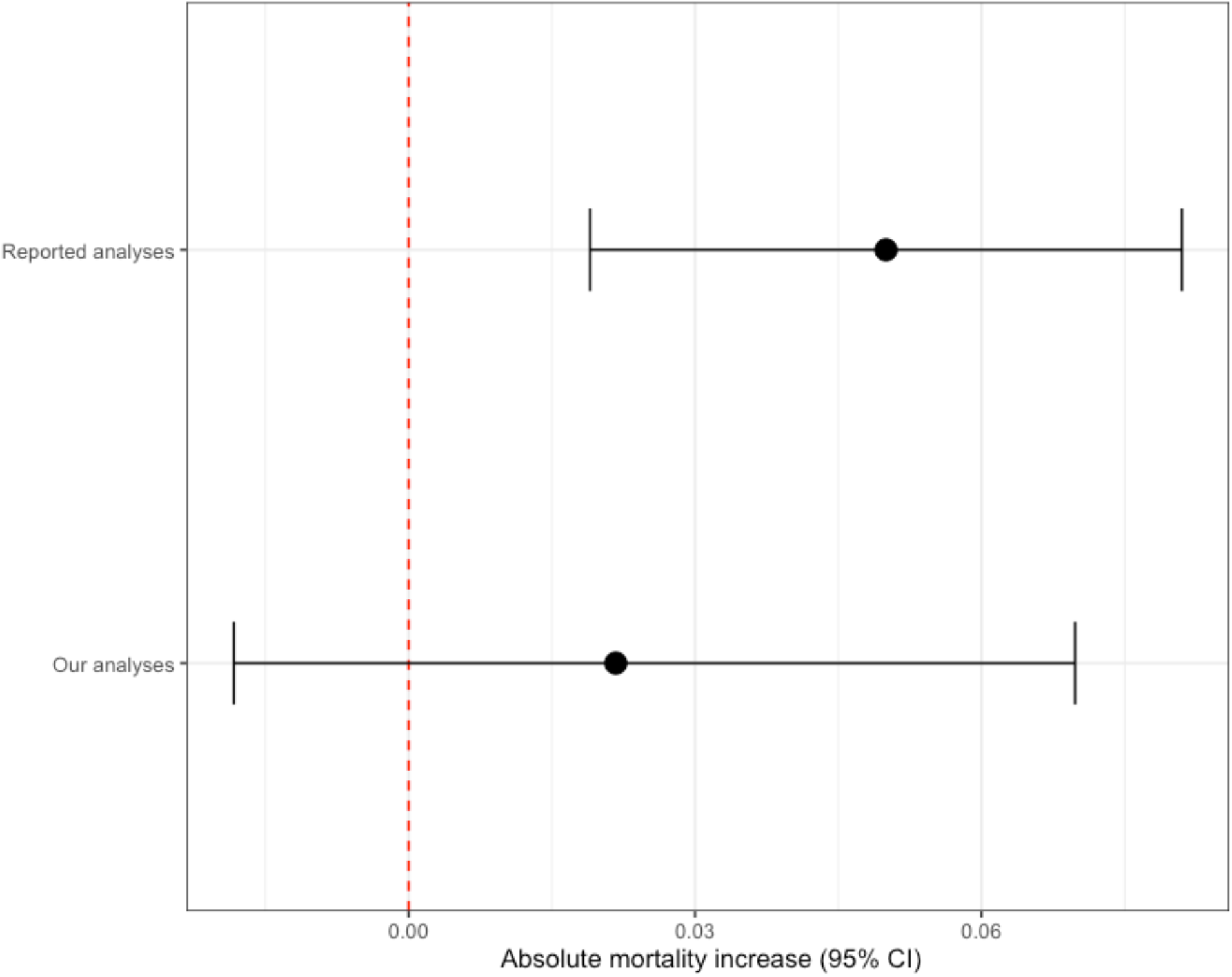
Absolute risk increase associated with piperacillin-tazobactam usage reported in the original manuscript, and with our estimates from summary level data. We recognise these are likely to be imprecise than estimates using individual level data.

## Discussion

Instrumental variable analyses can allow for causal inference when randomised trials are lacking, unethical, or impractical. Although in principle simple analytically, it is clear from our results above that they can be seriously biased when inappropriate control variables are included.

In this particular case, we think it is clear that the addition of metronidazole into the IV estimation is inappropriate, as it is a clinically relevant decision which is downstream of the instrumental variable. This can be seen as adjusting for a post-randomisation covariate in a randomised trial, which is explicitly warned against.

We recognise our simulations are based on one specified causal model which is an oversimplification of the true model. We do not state with certainty that the model we present represents truth, but it is certain that metronidazole usage is associated with more severe disease (i.e. there is unmeasured confounding), and that metronidazole usage is strongly associated with the IV. Using domain specific knowledge, our causal model seems plausible but we would encourage the authors to present further data supporting or refuting our analysis. We did ask the authors to provide analyses unadjusted for metronidazole which would help confirm if collider bias was occurring or whether our causal model was appropriate but they could not provide this data citing institutional regulations, so we are limited to our summary analysis, which does provide evidence that the unadjusted effect is much close to the null.

The authors do additionally present a secondary analysis not discussed above: whereby the exposure is the receipt of *either* piperacillin-tazobactam or metronidazole, as both of these have anaerobic activity. We note that the effect reported in this analysis is even stronger (12% increase in absolute mortality), and highly implausible given the null effect in the unadjusted analyses for piperacillin-tazobactam alone. This also raises challenges for IV analyses: the exposure is not a single drug, so we cannot state estimates represents the causal effect of increasing piperacillin-tazobactam or metronidazole individually: rather it represents the causal effect of “increased piperacillin-tazobactam and metronidazole usage” as a combined variable and cannot be interpreted as saying that giving more of either drug individually would lead to increased mortality. As with the primary analysis, it is likely that bias is generated via unmeasured confounding in this combined variable. It is worth thinking of the plausibility of situations whereby the causal effect would be predicted to almost double by changing the exposure definition to include a composite exposure that is less compellingly related to the IV (which was a shortage of piperacillin-tazobactam only).

The data reported from Chanderraj et al is therefore actually highly consistent with the recent ACORN trial^2^, and should be used as evidence that there is unlikely to be a large difference in mortality between piperacillin-tazobactam and cefepime based on these two studies. This is reassuring, and contrary to some of the reports surrounding this paper,^3^ clinicians should not favour cefepime simply based on this evidence.

We strongly suggest that consideration should be given to reporting the unadjusted results in the original manuscript.^1^ Currently, readers of the manuscript might assume that cefepime is a far superior agent to piperacillin-tazobactam which is not likely to be the case.

More broadly, we make the point that IV analyses are challenging to perform, and susceptible to biases like other observational studies. We recommend that authors, editors, and readers, interpret IV analyses cautiously, particularly when results appear to be contingent on a particular analytical set-up. The reason for our investigation of this issue was the identification that there was no convincing relationship between the shortage period and overall mortality but a 5% increase in mortality with piperacillin-tazobactam reported in the IV analysis. Although it is technically possible for IV estimates to be more precise and stronger than the raw outcome onto instrument relationship, this is highly unusual. It does not seem plausible that during a period in which there was almost complete changeover from piperacillin-tazobactam to cefepime, and in which there was no change in overall mortality, that cefepime could be associated with a 5% absolute reduction in mortality. The secondary analyses reporting an even larger 12% increase in mortality are equally implausible.

This highlights the value of examining each step in any analysis, and examining it for plausibility, and when implausible estimates are generated, examining the model inputs and outputs for potential bias and its causes.

## Conclusion

The reported estimates of benefit of cefepime in Chanderraj et al are likely due to collider bias due to conditioning on metronidazole. Corrected causal analyses identify a much-reduced effect. The authors should consider correcting the original study to include estimates unadjusted for metronidazole.

## Data Availability

All data presented is available, or in the supplement.

## REFERENCES

1. Chanderraj R, Admon AJ, He Y, et al. Mortality of Patients With Sepsis Administered Piperacillin-Tazobactam vs Cefepime. JAMA Intern Med [Internet] 2024; Available from: 10.1001/jamainternmed.2024.0581

2. Qian ET, Casey JD, Wright A, et al. Cefepime vs Piperacillin-Tazobactam in Adults Hospitalized With Acute Infection: The ACORN Randomized Clinical Trial. JAMA [Internet] 2023 [cited 2024 Jun 11];330(16):1557–67. Available from: https://jamanetwork.com/journals/jama/fullarticle/2810592

3. Dressler DD MSc MD. Cefepime vs. Piperacillin-Tazobactam for Sepsis: The Debate Continues [Internet]. 2024 [cited 2024 Jul 10];Available from: https://www.jwatch.org/na57489/2024/05/21/cefepime-vs-piperacillin-tazobactam-sepsis-debate

4. Greene WH. Econometric Analysis [Internet]. Prentice Hall; 2003. Available from: https://play.google.com/store/books/details?id=JJkWAQAAMAAJ

5. Deuchert E, Huber M. A cautionary tale about control variables in IV estimation. Oxf Bull Econ Stat [Internet] 2017;79(3):411–25. Available from: https://onlinelibrary.wiley.com/doi/10.1111/obes.12177

6. ICH E9 statistical principles for clinical trials - Scientific guideline [Internet]. [cited 2024 Jun 3];Available from: https://www.ema.europa.eu/en/ich-e9-statistical-principles-clinical-trials-scientific-guideline

7. GriUith GJ, Morris TT, Tudball M, et al. Collider bias undermines our understanding of COVID-19 disease risk and severity [Internet]. bioRxiv. 2020;Available from: 10.1101/2020.05.04.20090506

8. Sanderson E, Glymour MM, Holmes MV, et al. Mendelian randomization. Nature Reviews Methods Primers [Internet] 2022 [cited 2023 May 22];2(1):1–21. Available from: https://www.nature.com/articles/s43586-021-00092-5

9. Dobner J, Kaser S. Body mass index and the risk of infection - from underweight to obesity. Clin Microbiol Infect [Internet] 2018;24(1):24–8. Available from: 10.1016/j.cmi.2017.02.013

