## Supplement 1 for "Instrumental variable analysis: choice of control variables is critical and can lead to biased results"

### Instrumental Variable Analysis with Piperacillin-Tazobactam Shortage

Fergus Hamilton

#### Introduction

In this document, we perform an instrumental variable (IV) analysis using a simulated dataset to illustrate why adjusting for Metronidazole (MTZ) may lead to bias in estimating the effect of cefipime on mortality in the context of a piperacillin-tazobactam (pip-taz) shortage when MTZ is commonly used instead. We will also provide a Directed Acyclic Graph (DAG) to visually explain the relationships between the variables.

First, we generate a causal model where there is no effect of cefipime on mortality:

#### Directed Acyclic Graph (DAG)

Below is a DAG that represents the relationships between the variables in our analysis:

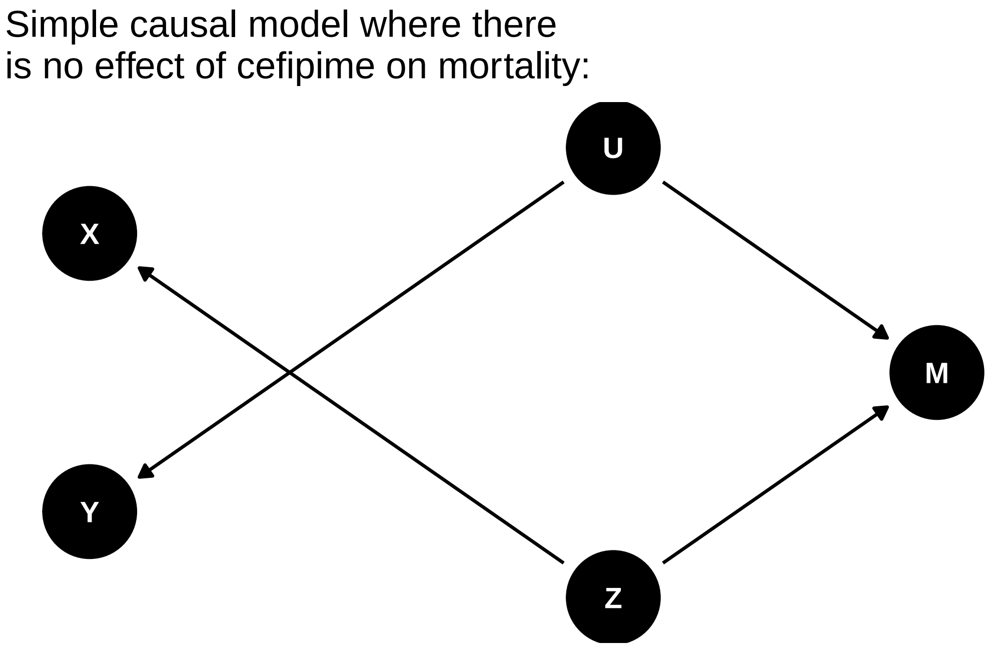


In this DAG:

- `Z is the instrument (Pip-Taz Shortage).`
- `X is the exposure (use of cefipime)`
- `M is Metronidazole use.`
- `Y is the outcome (e.g. mortality, or length of stay)`
- `U is an unmeasured confounder.`

Note, there is no effect of cefipime on mortality in this graph (there is no arrow between X and Y).

Adjusting for Metronidazole (M) in this context is problematic because M is influenced by both Z and U. By adjusting for M, we can introduce bias due to conditioning on a collider, which opens up a backdoor path that was otherwise blocked.

We can simulate this set up:

```
# Load necessary libraries
library(dplyr)
library(broom)
library(AER)  # For IV regression

# Set seed for reproducibility
set.seed(42)
```

```r
# Simulation parameters
n <- 50000
p <- 0.5  # Probability of Z (Pip-Taz Shortage)
q <- 0.5  # Probability of U (Confounder)

# Generate data
Z <- rbinom(n, 1, p)  # Pip-Taz Shortage (Instrument)
U <- rbinom(n, 1, q)  # Confounder

# Generate X based on Z
X <- rbinom(n, 1, plogis(0.5 + 3 * Z))

# Generate Metronidazole (M) based on Z and U with strong effects
M <- rbinom(n, 1, plogis(0.5 + 2 * Z + 2 * U))

# Generate survival probability (Y) based on U only with strong effects, and no causal effec
Y <- rbinom(n, 1, plogis(0.5 + 2 * U))

# Create DataFrame
data <- data.frame(Z = Z, U = U, X = X, M = M, Y = Y)

head(data)
```

```
  Z U X M Y
1 1 0 1 1 1
2 1 0 1 1 1
3 0 0 1 1 1
4 1 0 1 1 0
5 1 0 1 1 1
6 1 0 1 1 1
```

We can see that we have created a dataframe with 50,000 participants with the above causal paths. As can be seen in the above formula, there is no direct relationship between X (cefipime usage) and Y (mortality).

We can then run IV regression both unadjusted:

```r
# IV regression without adjusting for M
iv_model1 <- ivreg(Y ~ X | Z, data = data)
summary1 <- summary(iv_model1)
```

```r
# Display the results
print("IV Regression without adjusting for M")
```

```
[1] "IV Regression without adjusting for M"
```

```r
print(summary1)
```

```
Call:
ivreg(formula = Y ~ X | Z, data = data)

Residuals:
    Min      1Q  Median      3Q     Max 
-0.7811  0.2189  0.2280  0.2280  0.2280 

Coefficients:
             Estimate Std. Error t value Pr(>|t|)    
(Intercept)  0.781126   0.008906  87.706   <2e-16 ***
X           -0.009091   0.010925  -0.832    0.405    
---
Signif. codes:  0 '***' 0.001 '**' 0.01 '*' 0.05 '.' 0.1 ' ' 1

Residual standard error: 0.4183 on 49998 degrees of freedom
Multiple R-Squared: -2.006e-05, Adjusted R-squared: -4.007e-05 
Wald test: 0.6925 on 1 and 49998 DF,  p-value: 0.4053 
```

And adjusted for Metronidazole use:

```r
# IV regression adjusting for M
iv_model2 <- ivreg(Y ~ X + M | Z + M, data = data)
summary2 <- summary(iv_model2)


print("IV Regression adjusting for M")
```

```
[1] "IV Regression adjusting for M"
```

```r
print(summary2)
```

```
Call:
ivreg(formula = Y ~ X + M | Z + M, data = data)

Residuals:
    Min      1Q  Median      3Q     Max
-0.8456  0.1544  0.2251  0.2251  0.3399

Coefficients:
             Estimate Std. Error t value Pr(>|t|)
(Intercept)  0.730801   0.009226  79.214  < 2e-16 ***
X           -0.070727   0.011328  -6.243 4.32e-10 ***
M            0.114794   0.005700  20.138  < 2e-16 ***
---
Signif. codes:  0 '***' 0.001 '**' 0.01 '*' 0.05 '.' 0.1 ' ' 1

Residual standard error: 0.4174 on 49997 degrees of freedom
Multiple R-Squared: 0.004534,   Adjusted R-squared: 0.004495
Wald test: 203.1 on 2 and 49997 DF,  p-value: < 2.2e-16
```

We can also plot these estimates:

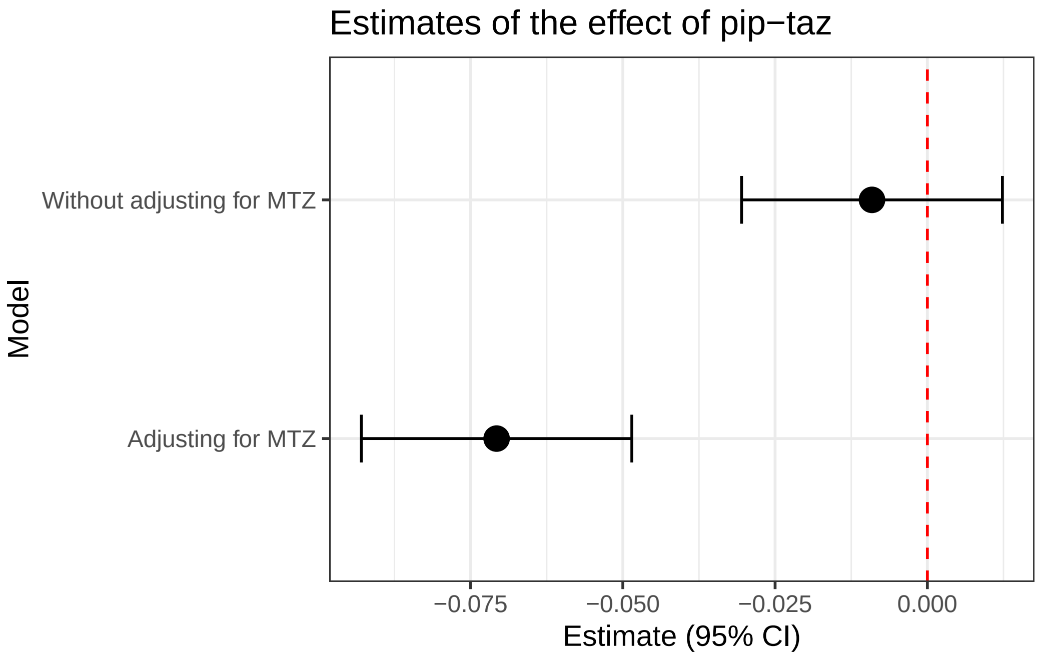


As can be seen; adjusting for M leads to a strong estimated effect of X onto Z. This suggests - incorrectly for this causal model - that cefipime usage is associated with reduced mortality.

Adjusting for metronidazole (M) in the analysis introduces bias because M is affected by both the instrument (Z) and the confounder (U). When we condition on MTZ, we inadvertently open a backdoor path from Z to Y through U, which can introduce spurious associations and bias the estimate of the effect of X on Y.

To understand this, think of metronidazole as a marker for more severe illness. Patients who are more severely ill are more likely to receive metronidazole. In our DAG, (M) is also influenced by the instrument (Z, pip-taz shortage) because during a shortage, doctors prescribe MTZ more often. This is shown in the paper. MTZ is influenced by the confounder (U), which represents the severity of illness.

When we adjust for M, we are essentially saying, "Let's compare patients who received metronidazole to those who didn't, within each level of metronidazole use." However, since metronidazole use is driven by both the shortage and the severity of illness, this comparison mixes up the effects of the shortage and the illness severity.

In simpler terms:

• Without adjusting for metronidazole: We are comparing mortality rates between those who had used cefipime and those who didn't, while ignoring metronidazole use.

• Adjusting for metronidazole: We are comparing mortality rates between those who had access to pip-taz and those who didn't, within the groups of those who used metronidazole and those who didn't. This comparison is flawed because metronidazole use itself is influenced by how sick patients are, which is not directly related to the pip-taz shortage.

Therefore, adjusting for M breaks the assumption that the instrument (Z) only affects the outcome (Y) through the exposure (cefipime usage), leading to biased estimates.

Conclusion

Our analysis illustrates that adjusting for colliders such as metronidazole can introduce bias in instrumental variable analyses. In the context of a piperacillin-tazobactam shortage where MTZ is commonly used as an alternative, it is crucial to avoid adjusting for MTZ to maintain the validity of the IV assumptions.

#### Supplement: what happens when you run the analyses in MTZ users and non-MTZ users?

Running the analysis stratified also leads to selection bias:

```r
# Create DataFrame
data <- data.frame(Z = Z, U = U, X = X, M = M, Y = Y)

mtz_df <- data %>%
  filter(M == 1)

nomtz_df <- data %>%
  filter(M == 0)

# IV regression users
iv_model1 <- ivreg(Y ~ X | Z, data = mtz_df)
summary1 <- summary(iv_model1)

# IV regression non-users
iv_model2 <- ivreg(Y ~ X  | Z , data = nomtz_df)
summary2 <- summary(iv_model2)

# Extract coefficients and confidence intervals
estimates <- data.frame(
  model = c("Metronidazole users", "Metronidazole non-users"),
  estimate = c(coef(summary1)[2, 1], coef(summary2)[2, 1]),
  conf.low = c(confint(iv_model1)[2, 1], confint(iv_model2)[2, 1]),
  conf.high = c(confint(iv_model1)[2, 2], confint(iv_model2)[2, 2])
)

# Create forest plot
ggplot(estimates, aes(x = model, y = estimate)) +
  geom_point(size = 4) +
  geom_hline(aes(yintercept = 0), col = "red", lty = "dashed") +
  geom_errorbar(aes(ymin = conf.low, ymax = conf.high), width = 0.2) +
  labs(title = "Estimates of the effect of pip-taz: stratified",
       x = "Model",
       y = "Estimate (95% CI)") +
  theme_bw() +
  coord_flip()
```

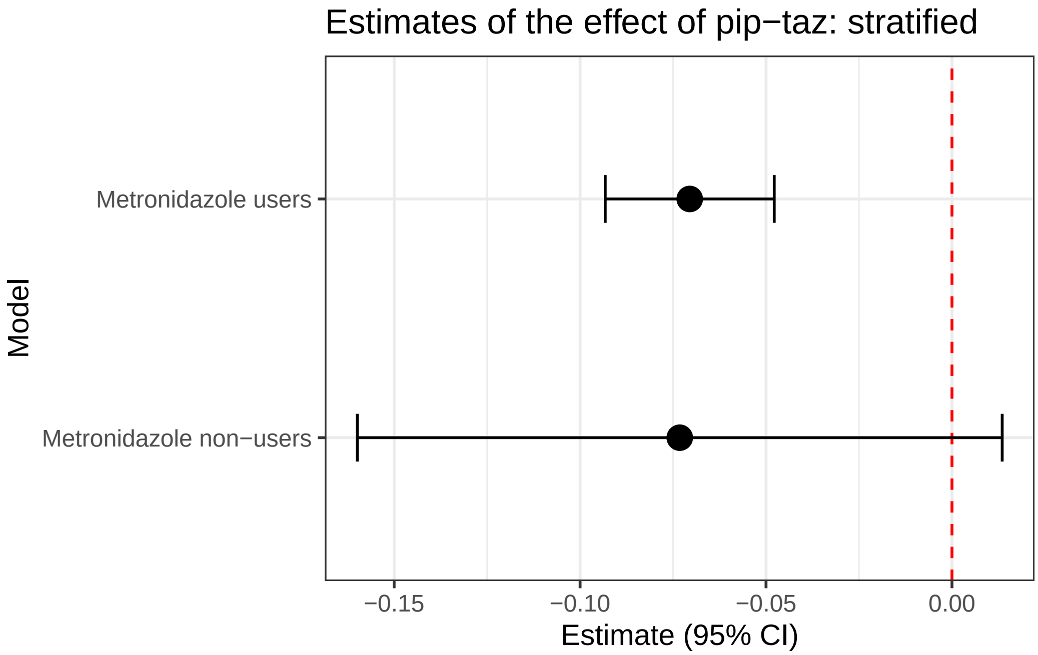


Now, to run where X -> M instead of Z -> M

#### Directed Acyclic Graph (DAG)

Below is a DAG that represents the relationships between the variables in our analysis:

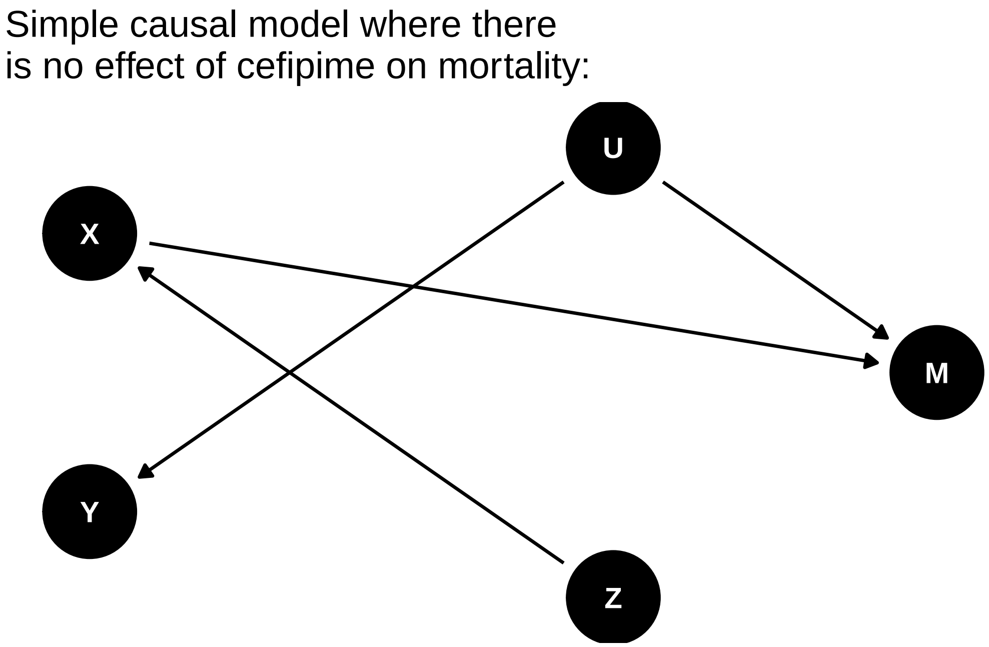


In this DAG:

- `Z is the instrument (Pip-Taz Shortage).`
- `X is the exposure (use of cefipime)`
- `M is Metronidazole use.`
- `Y is the outcome (e.g. mortality, or length of stay)`
- `U is an unmeasured confounder.`

Note, there is no effect of cefipime on mortality in this graph (there is no arrow between X and Y).

Adjusting for Metronidazole (M) in this context is problematic because M is influenced by both Z and U. By adjusting for M, we can introduce bias due to conditioning on a collider, which opens up a backdoor path that was otherwise blocked.

We can simulate this set up:

```
# Load necessary libraries
library(dplyr)
library(broom)
library(AER)  # For IV regression

# Set seed for reproducibility
set.seed(42)
```

```r
# Simulation parameters
n <- 50000
p <- 0.5  # Probability of Z (Pip-Taz Shortage)
q <- 0.5  # Probability of U (Confounder)

# Generate data
Z <- rbinom(n, 1, p)  # Pip-Taz Shortage (Instrument)
U <- rbinom(n, 1, q)  # Confounder

# Generate X based on Z
X <- rbinom(n, 1, plogis(0.5 + 3 * Z ))

# Generate Metronidazole (M) based on Z and U with strong effects
M <- rbinom(n, 1, plogis(0.5 + 2 * X + 2 * U))

# Generate survival probability (Y) based on U only with strong effects, and no causal effec
Y <- rbinom(n, 1, plogis(0.5 + 2 * U))

# Create DataFrame
data <- data.frame(Z = Z, U = U, X = X, M = M, Y = Y)

head(data)
```

```
  Z U X M Y
1 1 0 1 1 1
2 1 0 1 1 1
3 0 0 1 1 1
4 1 0 1 1 0
5 1 0 1 1 1
6 1 0 1 1 1
```

We can see that we have created a dataframe with 50,000 participants with the above causal paths. As can be seen in the above formula, there is no direct relationship between X (cefipime usage) and Y (mortality).

We can then run IV regression both unadjusted:

```r
# IV regression without adjusting for M
iv_model1 <- ivreg(Y ~ X | Z, data = data)
summary1 <- summary(iv_model1)
```

```r
# Display the results
print("IV Regression without adjusting for M")
```

```
[1] "IV Regression without adjusting for M"
```

```r
print(summary1)
```

```
Call:
ivreg(formula = Y ~ X | Z, data = data)

Residuals:
    Min      1Q  Median      3Q     Max 
-0.7811  0.2189  0.2280  0.2280  0.2280 

Coefficients:
             Estimate Std. Error t value Pr(>|t|)    
(Intercept)  0.781126   0.008906  87.706   <2e-16 ***
X           -0.009091   0.010925  -0.832    0.405    
---
Signif. codes:  0 '***' 0.001 '**' 0.01 '*' 0.05 '.' 0.1 ' ' 1

Residual standard error: 0.4183 on 49998 degrees of freedom
Multiple R-Squared: -2.006e-05, Adjusted R-squared: -4.007e-05 
Wald test: 0.6925 on 1 and 49998 DF,  p-value: 0.4053 
```

And adjusted for Metronidazole use:

```r
# IV regression adjusting for M
iv_model2 <- ivreg(Y ~ X + M | Z + M, data = data)
summary2 <- summary(iv_model2)


print("IV Regression adjusting for M")
```

```
[1] "IV Regression adjusting for M"
```

```r
print(summary2)
```

```
Call:
ivreg(formula = Y ~ X + M | Z + M, data = data)

Residuals:
    Min      1Q  Median      3Q     Max 
-0.8082  0.1918  0.2231  0.2231  0.3402 

Coefficients:
             Estimate Std. Error t value Pr(>|t|)    
(Intercept)  0.691070   0.008287  83.393  < 2e-16 ***
X           -0.031315   0.011713  -2.674  0.00751 ** 
M            0.117106   0.008345  14.032  < 2e-16 ***
---
Signif. codes:  0 '***' 0.001 '**' 0.01 '*' 0.05 '.' 0.1 ' ' 1

Residual standard error: 0.4173 on 49997 degrees of freedom
Multiple R-Squared: 0.005038,	Adjusted R-squared: 0.004998 
Wald test: 118.7 on 2 and 49997 DF,  p-value: < 2.2e-16 
```

We can also plot these estimates:

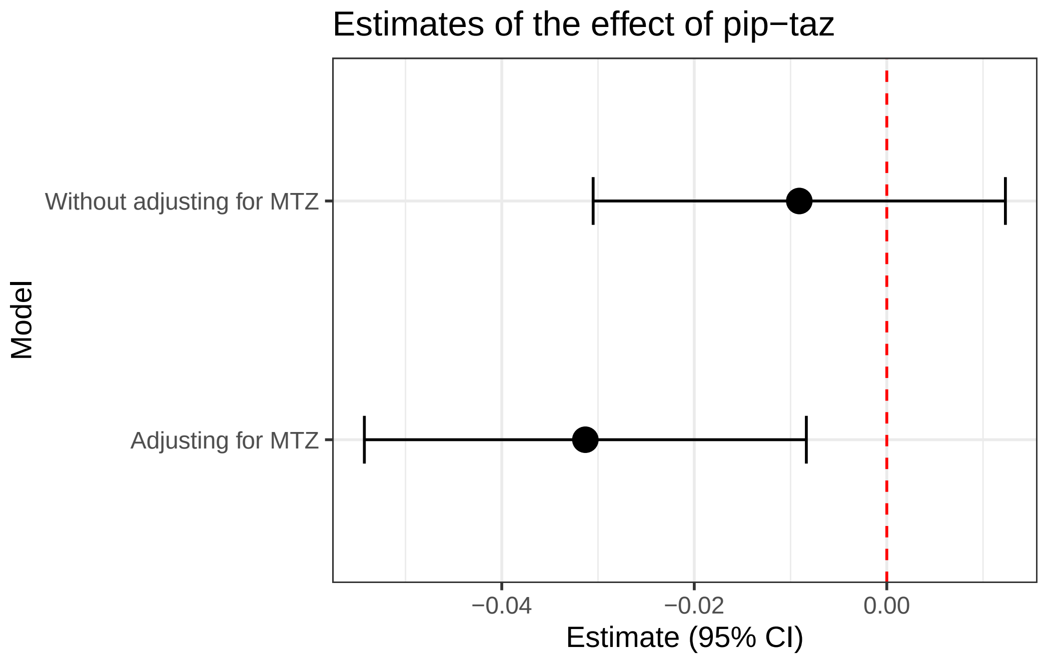
