## Supplement 2 for "Instrumental variable analysis: choice of control variables is critical and can lead to biased results"

Supplemental Note: Comparing p-values in TSLS and IV Analysis

Fergus Hamilton

### Supplemental Note 1

Addendum: IV-analysis using the raw data from the paper. For completeness, we estimated the TSLS estimate from the raw presented data. This will not match exactly but should be close to an unadjusted, raw TSLS estimate.

#### Raw Data from the Paper

- **First Stage (IV-Exposure Effect)**:
  - Effect of IV (Within shortage) on exposure (pip-taz treatment probability)
  - $\beta=-0.659$
  - 95% Confidence Interval: $\left[ -0.68,-0.64 \right]$
  - p-value: $<0.001$
  - F-statistic: 4535
- **Mortality Data**:
  - Number of patients outside shortage: 5460
  - Number of patients within shortage: 2109
  - Number of deaths outside shortage: 1137
  - Number of deaths within shortage: 417

#### IV-Exposure Effect (First Stage)

From the first stage data:

$$\pi_{1}=-0.659$$

#### Logistic Regression for IV-Outcome Effect

Using logistic regression to estimate the IV-outcome effect and its standard error.

##### Logistic Regression Results:

- Coefficient ($\alpha_{1}$): -0.0650
- Standard Error ($\text{SE}\left( \hat{\alpha}_{1} \right)$): 0.0640

#### TSLS Estimate

Using the IV estimates from the first stage and the logistic regression:

$$\alpha_{1}=-0.0650$$

$$\beta_{IV}=\frac{\alpha_{1}}{\pi_{1}}=\frac{-0.0650}{-0.659}=0.0987$$

#### Standard Error of the TSLS Estimate

Given the standard errors: - $\text{SE}\left( \hat{\alpha}_{1} \right)=0.0640$ - $\text{SE}\left( \hat{\pi}_{1} \right)=0.0102$

The standard error of the TSLS estimate is:

$$\text{SE}\left( \hat{\beta}_{IV} \right)=\sqrt{\frac{\left( 0.0640 \right)^{2}}{\left( -0.659 \right)^{2}}+\left( 0.0987 \right)^{2}\cdot\frac{\left( 0.0102 \right)^{2}}{\left( -0.659 \right)^{2}}}$$

$$\text{SE}\left( \hat{\beta}_{IV} \right)=\sqrt{0.00937744+0.00023125}\approx0.0972$$

#### 95% Confidence Interval for TSLS Estimate

The 95% Confidence Interval is:

$$\text{CI}=0.0987\pm1.96\times0.0972=\left[ 0.0987-0.1905,0.0987+0.1905 \right]=\left[ -0.0918,0.2891 \right]$$

#### Reporting TSLS as Odds Ratio and Absolute Risk Increase

The TSLS estimate in terms of odds ratio (OR) can be calculated as:

$$\text{OR}=e^{\beta_{IV}}=e^{0.0987}\approx1.1037$$

To estimate the absolute risk increase, we convert the OR to risk difference:

Given the baseline mortality rate outside the shortage period is 20.8%, we can estimate the absolute risk increase.

$$\text{Absolute Risk Increase}=\text{Baseline Risk}\times\left( \text{OR}-1 \right)$$

$$\text{Absolute Risk Increase}=0.208\times\left( 1.1037-1 \right)\approx0.0217\text{ or 2.17\%}$$

#### 95% Confidence Interval for Odds Ratio

For the 95% confidence interval of the OR, we use the confidence limits of the TSLS estimate:

$$\text{OR Lower CI}=e^{-0.0918}\approx0.9123$$

$$\text{OR Upper CI}=e^{0.2891}\approx1.3355$$

#### 95% Confidence Interval for Absolute Risk Increase

For the absolute risk increase, we calculate the bounds as follows:

$$\text{Absolute Risk Increase Lower CI}=0.208\times\left( 0.9123-1 \right)\approx-0.0183\text{ or -1.83\%}$$

$$\text{Absolute Risk Increase Upper CI}=0.208\times\left( 1.3355-1 \right)\approx0.0698\text{ or 6.98\%}$$
